# Uncovering Gender Dimensions in Antimicrobial Resistance: A 10-Year Study of Clinical Bacterial Isolates in Uganda

**DOI:** 10.1101/2025.04.09.25325544

**Authors:** Joel Bazira, Nakato Constance Nakimuli, Pauline Petra Nalumaga, Brenda Nakazibwe, Abel. W. Walekhwa, Kawuma Simon, Hope Derick, Iramiot Jacob, Lawrence Mugisha

## Abstract

**Background:** Antimicrobial resistance (AMR) and multidrug resistance (MDR) are escalating global health threats, particularly in low- and middle-income countries (LMICs). Understanding gender-specific resistance patterns is essential for inclusive antimicrobial stewardship and gender-targeted interventions.

**Objective:** To investigate gender-specific trends in AMR and MDR among clinical bacterial isolates collected from Mbarara Regional Referral Hospital, Uganda, between a 10-year period (2014–2024).

**Methods:** A total of 4,170 non-duplicate clinical isolates subjected to antimicrobial susceptibility testing (AST) were retrospectively analyzed. Gender-specific resistance burdens were calculated and compared using the Wilcoxon Signed-Rank Test, Chi-square, and Fisher’s Exact Test. All analyses were performed in Python (Google Colab).

**Results:** Among all isolates, 92.7% were resistant to at least one antibiotic. Female patient isolates accounted for 59.8% of all resistant organisms and exhibited significantly higher resistance rates across multiple classes, including beta-lactams, fluoroquinolones, and aminoglycosides (Wilcoxon W = 991.000, p = 0.00009). Multidrug resistance was significantly more common in female-derived *Escherichia coli* and *Staphylococcus aureus* isolates (p = 0.0133 and p = 0.002, respectively), while MDR *Pseudomonas aeruginosa* was more prevalent in males.

**Conclusion:** This study reveals distinct gender disparities in AMR and MDR patterns, underscoring the need to integrate sex-disaggregated indicators into AMR surveillance and stewardship efforts. These findings advocate for gender-responsive antimicrobial policies to ensure equitable public health outcomes.

## INTRODUCTION

Antimicrobial resistance (AMR) is one of the most pressing public health challenges of the 21st century, contributing to approximately 1.27 million deaths and disproportionately affecting low- and middle-income countries (LMICs) where diagnostic capacity and antimicrobial stewardship infrastructure remain limited (1). In Uganda and across Sub-Saharan Africa, AMR has significantly contributed to mortality associated with bloodstream and urinary tract infections, particularly those caused by *Escherichia coli* and *Klebsiella pneumoniae* (2, 3).

Multidrug resistance (MDR)—defined as resistance to at least one agent in three or more antimicrobial classes exacerbates the challenge by narrowing therapeutic options and increasing treatment failures, costs, and mortality (4). While surveillance systems increasingly monitor AMR trends by bacterial species and drug class, few studies have examined resistance trends through a gender lens, especially in LMIC settings.

Biological, behavioral, and structural factors contribute to sex-based differences in infection risk and antibiotic exposure. Women, for instance, experience more frequent urinary tract infections and are more likely to receive empiric antibiotic treatment for reproductive or urogenital symptoms, potentially leading to greater antimicrobial exposure and resistance development (5, 6). Despite this, gender remains a largely neglected variable in microbiological surveillance systems and antimicrobial resistance research in Africa (7).

Evidence from high-income settings has demonstrated sex-based AMR differences—for example, higher aminopenicillin resistance in women and higher methicillin resistance in men (8). However, comparable analyses from Sub-Saharan Africa are scarce. Systematic reviews have highlighted the urgent need for research exploring the intersection of gender and AMR in African contexts, where health system inequities may amplify underlying disparities (6). Therefore, this study aimed to determine gender-specific trends in AMR and MDR and to generate evidence to inform equitable surveillance strategies and gender-sensitive antimicrobial stewardship efforts.

## Methodology

### Study area

This was a retrospective, cross-sectional study done at Mbarara Regional Referral Hospital (MRRH) located in Mbarara, western Uganda, about 260 kilometers from Kampala capital city, and is also a teaching hospital for Mbarara University of Science and Technology. The hospital provides a wide range of health services through its departments of pediatrics, obstetrics and gynecology, internal medicine, surgery, cancer unit, emergency and critical care, imaging, pathology, laboratories, and outpatient departments. The hospital is a referral hospital for the Ankole sub-region, neighboring districts; specifically Mbarara, Bushenyi, Ntungamo, Kiruhura, Ibanda, and Isingiro, and even surrounding countries, such as Rwanda and Tanzania (9).

The study reviewed AST records from January 2014 to December 2024. Laboratory and clinical data were obtained from the hospital’s microbiology computer database. The 4,170 non-redundant bacterial isolates had each been matched with patient sex, organism type, specimen type, and corresponding antibiotic susceptibility results. The specimens were collected from a broad range of clinical sites such as urine, pus, blood, sputum, tracheal aspirate, wound swab, pleural fluid, and ascitic tap. Only cases with full AST profiles and recorded patient sex were included in the study.

### Culture methods and susceptibility testing

The collected samples were inoculated with a 0.01-mL loop on blood, chocolate, and MacConkey agar (HI MEDIA, INDIA) and incubated at 37 °C for 24 hours. Detected pathogens in significant amounts were identified according to phenotypical characteristics (colonial characteristics on the culture media, biochemical tests such as gram staining, catalase, oxidase, triple sugar iron, and sulfur indole motility) (10). AST was performed using the Kirby–Bauer disk diffusion method on Mueller–Hinton agar and interpreted according to Clinical and Laboratory Standards Institute (CLSI) guidelines.

Bacterial pathogens under investigation in this study were *Staphylococcus aureus, Escherichia coli, Klebsiella pneumoniae, Klebsiella species, Streptococcus pneumoniae, Salmonella species, Citrobacter species, Pseudomonas aeruginosa, Pseudomonas* species*, Enterobacter* species*, Acinetobacter* species*, Enterococcus* species*, and Streptococcus* species. Organism–antibiotic combinations with at least 30 isolates only were investigated to ensure statistical reliability and minimize sampling bias.

Antibiotics tested were grouped according to pharmacologic class. These included penicillins (amoxicillin, amoxicillin/clavulanic acid, ampicillin, penicillin, oxacillin, and piperacillin), cephalosporins (cefaclor, cefazolin, cefepime, cefixime, cefotaxime, cefovecin, cefoxitin, ceftriaxone, and cefuroxime), carbapenems (imipenem), monobactams (aztreonam), aminoglycosides (amikacin, gentamicin), fluoroquinolones (ciprofloxacin, levofloxacin), macrolides (azithromycin, erythromycin), lincosamides (clindamycin), tetracyclines (doxycycline, tetracycline), sulfonamides (sulfamethoxazole), phenicols (chloramphenicol), oxazolidinones (linezolid), glycopeptides (vancomycin), and nitrofurans (nitrofurantoin). Control strains used were *S. aureus* ATCC 25923 and *Klebsiella pneumoniae* ATCC 700603.

AMR is defined as resistance against a single or multiple antibiotics (11). Multidrug resistance (MDR) was defined as resistance against single or multiple agents in three or more antibiotic classes (12). Gender-based analysis used the total number of isolates tested for every bacterium–antibiotic combination as the denominator.

### Data analysis

Data was analyzed with Python 3.10 in Google Colaboratory. The resistance rates among male and female patients were compared using the Wilcoxon Signed-Rank Test for 99 bacteria–antibiotic pairs. Chi-square and Fisher’s Exact Tests were applied to analyze gender differences in MDR distribution at a significance level of p < 0.05.

## RESULTS

### Distribution of Clinical Specimens by Gender

A total of **4,170** clinical specimens for which antimicrobial susceptibility testing (AST) was performed between 2014 and 2024 were analyzed in this study. Of these, 59% were female patients and 41% were male patients. The most common specimens were urine samples, which accounted for over one-third of all specimens with 62.6% from female patients and 37.4% from male patients as shown in Table 1. The second most commonly collected specimen type was blood culture, with a fairly even gender split of 46.6% from females and 53.4% from males. Sputum samples were sampled more among males (55.6%%) than females (44.4%). Pus swabs, and infection episodes of soft tissues, were proportionally equally split in cases involving males and females each accounting for nearly half the total of 377 obtained.

**Table 1:** Gender distribution of clinical specimens.

| Specimen Type | Female (n) | Male (n) | Total (n) | % Female | % Male |
| --- | --- | --- | --- | --- | --- |
| Ascitic Tap | 11 | 10 | 21 | 52.4% | 47.6% |
| Blood Culture | 417 | 478 | 895 | 46.6% | 53.4% |
| Ear Swab | 45 | 50 | 95 | 47.4% | 52.6% |
| High Vaginal Swab (HVS) | 518 | 0 | 518 | 100.0% | 0.0% |
| Myocardial Infarct Tissue | 47 | 28 | 75 | 62.7% | 37.3% |
| Pleural Fluid | 13 | 20 | 33 | 39.4% | 60.6% |
| Pus Swab | 190 | 187 | 377 | 50.4% | 49.6% |
| Sputum | 228 | 285 | 513 | 44.4% | 55.6% |
| Stool | 15 | 13 | 28 | 53.6% | 46.4% |
| Throat Swab | 29 | 27 | 56 | 51.8% | 48.2% |
| Tracheal Aspirate | 28 | 51 | 79 | 35.4% | 64.6% |
| Urine | 852 | 509 | 1,361 | 62.6% | 37.4% |
| Wound Swab | 68 | 51 | 119 | 57.1% | 42.9% |
| **Total** | **2,461** | **1,709** | **4,170** | **59.0%** | **41.0%** |

*Table 1: Gender distribution of clinical specimens.*
| Specimen Type | Female (n) | Male (n) | Total (n) | % Female | % Male |
| --- | --- | --- | --- | --- | --- |
| Ascitic Tap | 11 | 10 | 21 | 52.4% | 47.6% |
| Blood Culture | 417 | 478 | 895 | 46.6% | 53.4% |
| Ear Swab | 45 | 50 | 95 | 47.4% | 52.6% |
| High Vaginal Swab (HVS) | 518 | 0 | 518 | 100.0% | 0.0% |
| Myocardial Infarct Tissue | 47 | 28 | 75 | 62.7% | 37.3% |
| Pleural Fluid | 13 | 20 | 33 | 39.4% | 60.6% |
| Pus Swab | 190 | 187 | 377 | 50.4% | 49.6% |
| Sputum | 228 | 285 | 513 | 44.4% | 55.6% |
| Stool | 15 | 13 | 28 | 53.6% | 46.4% |
| Throat Swab | 29 | 27 | 56 | 51.8% | 48.2% |
| Tracheal Aspirate | 28 | 51 | 79 | 35.4% | 64.6% |
| Urine | 852 | 509 | 1,361 | 62.6% | 37.4% |
| Wound Swab | 68 | 51 | 119 | 57.1% | 42.9% |
| **Total** | **2,461** | **1,709** | **4,170** | **59.0%** | **41.0%** |

Tissue from myocardial infarction was taken more frequently in females, who provided approximately 62.7%. At the same time, pleural fluid was more frequently taken in males (60.6%) while tracheal aspirates, another respiratory sample, were taken predominantly in males (64.6%). Wound swabs stool samples and ascitic taps were almost equally distributed between male and female patients.

### Distribution of Antibiotic-Resistant Isolates by Gender

Among the isolates, 3,867 (92.73%), were resistant to at least one antibiotic. Resistant isolates were more frequently detected in female patients, accounting for 59.8%, while 40.2% were from male patients, as shown in Figure 1.

**Figure 1:**
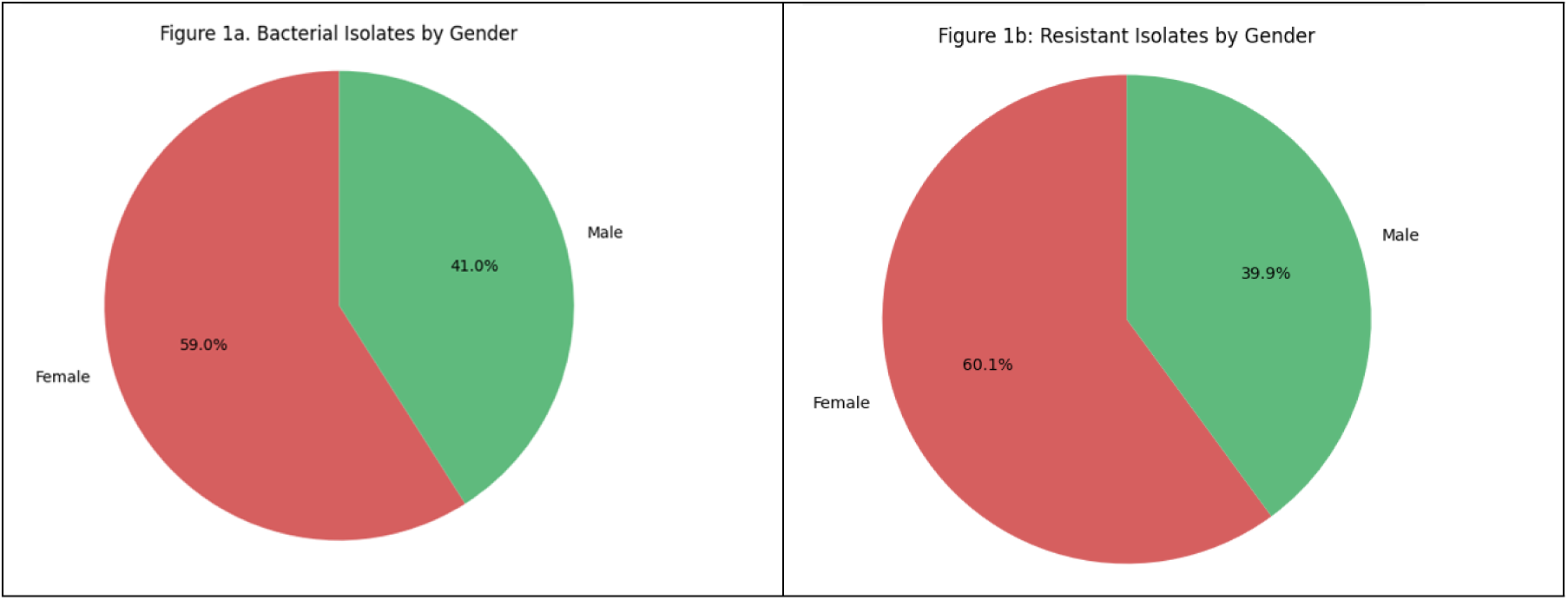
(a) Distribution of Bacterial Isolates by Gender and (b) Distribution of Antibiotic-Resistant Isolates by Gender.

Resistance to penicillins, macrolides, tetracyclines, and sulfonamides was higher in female isolates, while resistance to aminoglycosides, glycopeptides, and fluoroquinolones showed relatively smaller differences between genders while Carbapenem resistance remained lower in both groups (figure 2).

**Figure 2:**
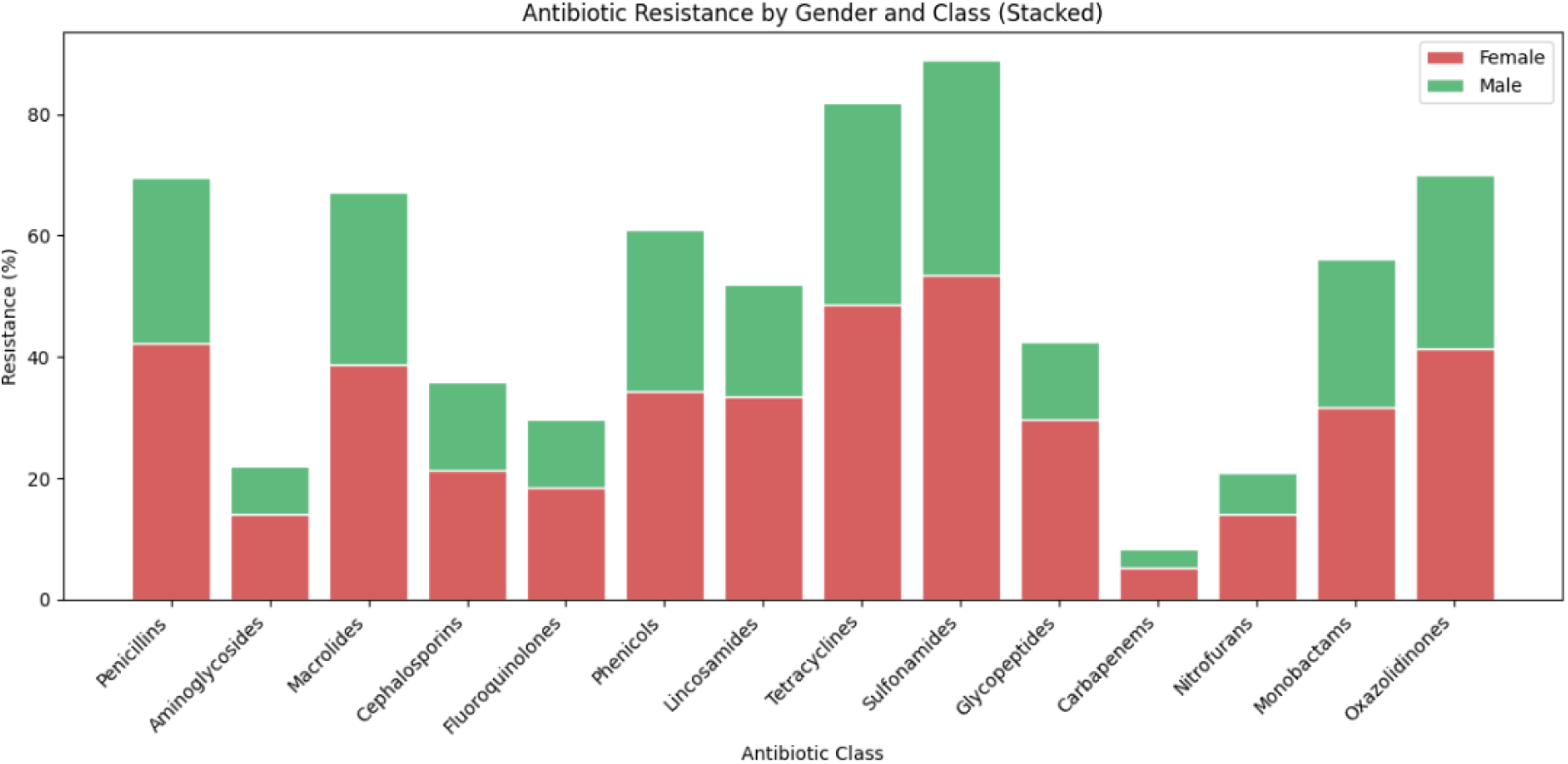
Antibiotic Resistance by Gender and Class.

### Antibiotic Resistance Patterns among Gram-Positive and Gram-Negative Bacteria

Resistance patterns to antibiotics varied among Gram-positive and Gram-negative bacterial isolates (Figures 3 and 4). Among Gram-positive organisms, resistance to sulfonamide and tetracycline was widespread. *Streptococcus* species were completely resistant to sulfonamides and over 94 % resistant to tetracyclines, while *Staphylococcus aureus* showed over 82 % resistance to tetracyclines and over 73 % to penicillins. Resistance to macrolides was also present, and it had gone up to 87.5 % in *Streptococcus* and nearly 69 % in *Staphylococcus aureus*. Resistance was lower to glycopeptides and cephalosporins, with *Enterococcus* species showing only 18 % resistance to glycopeptides and nearly 32 percent to cephalosporins.

**Figure 3:**
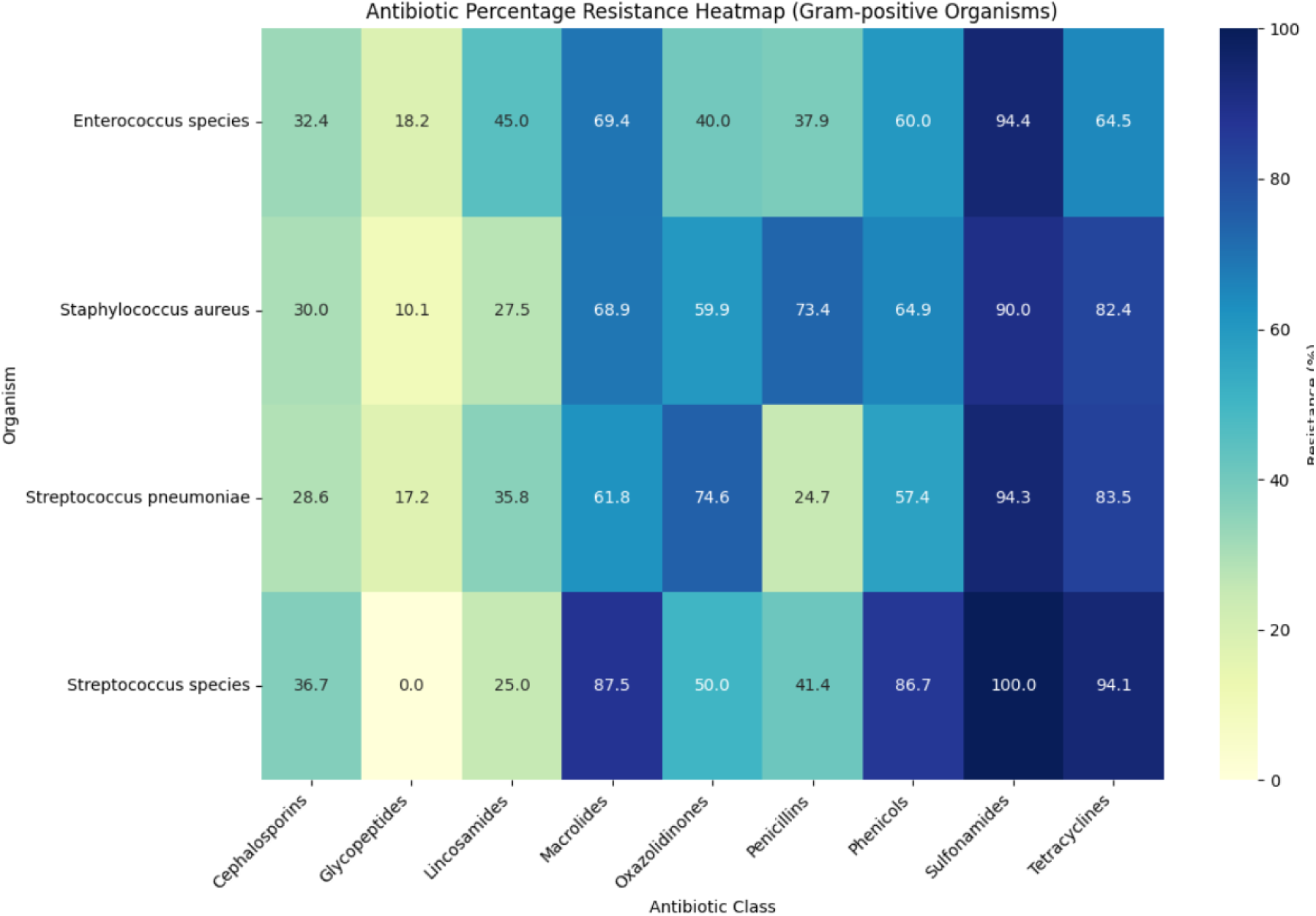
Antibiotic resistance trends in Gram-positive bacterial isolates.

**Figure 4:**
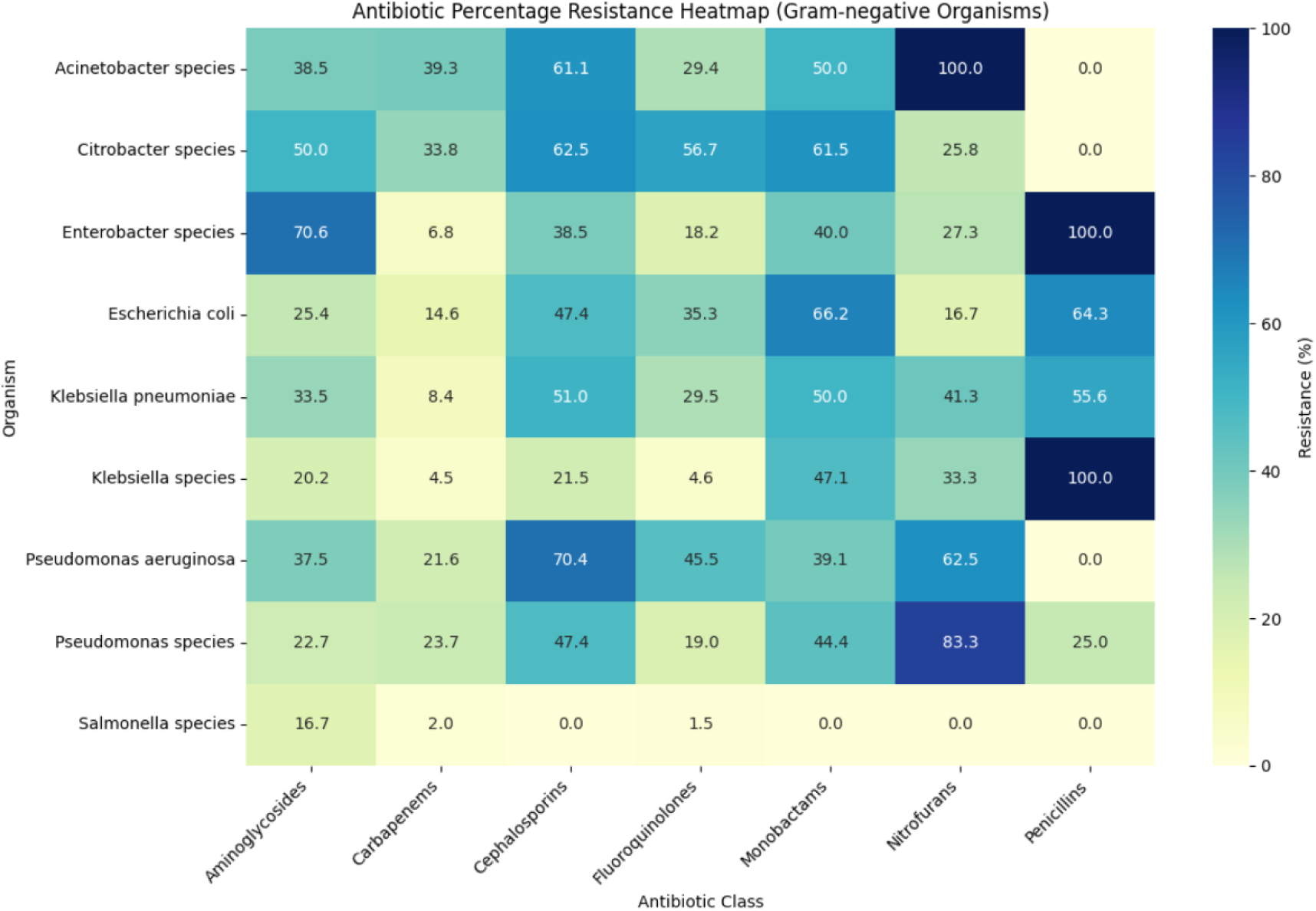
Antibiotic resistance patterns in Gram-negative bacterial isolates.

**Figure 5:**
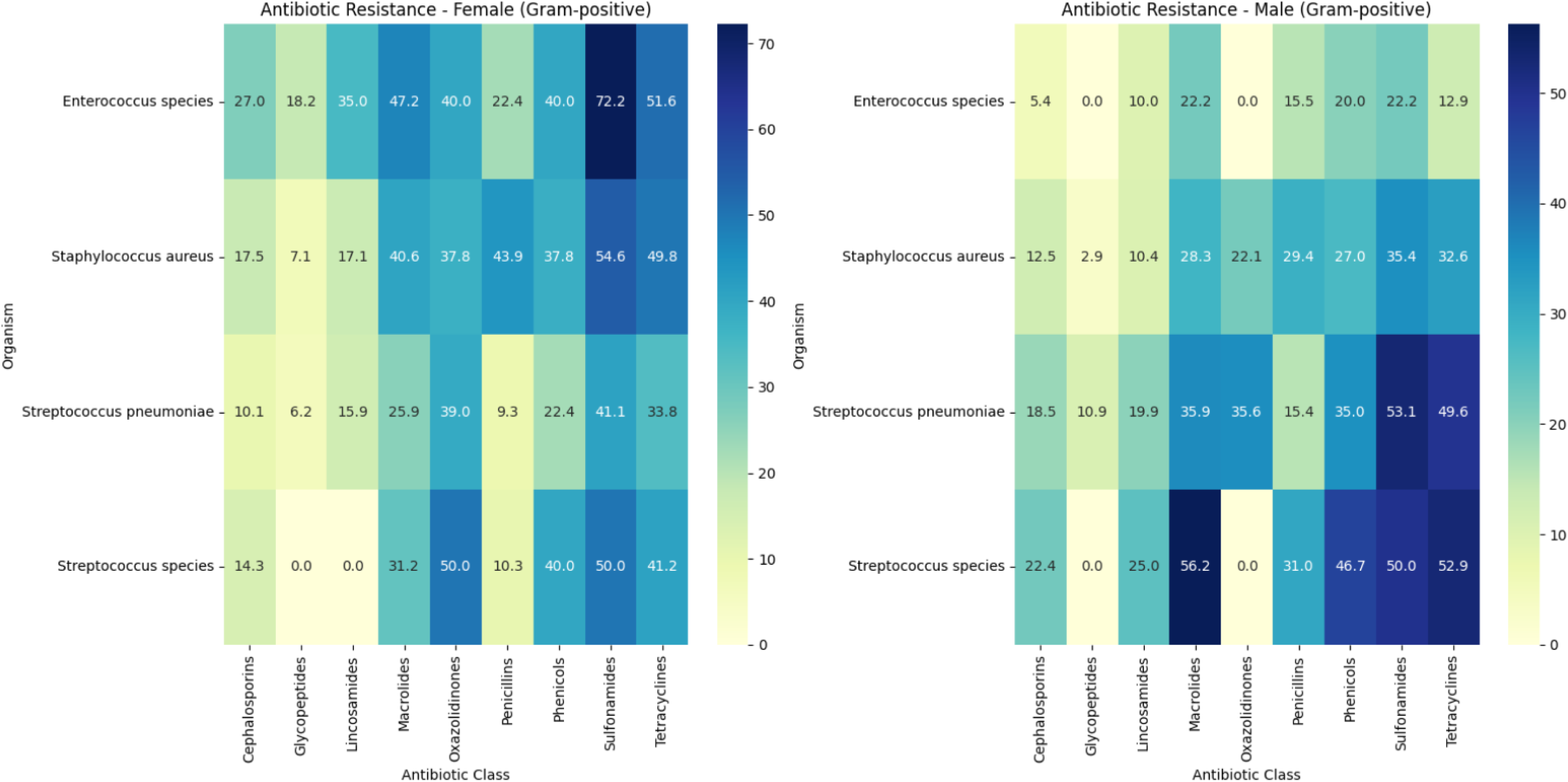
Gender-Specific Resistance in Gram-Positive Organisms

Among Gram-negative bacteria, carbapenem resistance was highest in *Acinetobacter* species with over 61 % of the isolates being resistant. Fluoroquinolone resistance was also high, with 83 % of *Pseudomonas* species being resistant. The greatest aminoglycoside resistance was exhibited by *Enterobacter* species at over 70 %, and *Escherichia coli* exhibited approximately 47 percent cephalosporin resistance and over 64 % to nitrofurans. The least resistance was shown by *Salmonella* species at a mere 16.7 % for aminoglycosides and almost no resistance against other types of antibiotics.

### Gender-Specific Trends in Antibiotic Resistance

Trends in antimicrobial resistance varied among bacterial isolates obtained from male and female patients, showing distinct differences between Gram-positive and Gram-negative bacteria.

### Gram-Positive Bacteria

Amongst the *Streptococcus pneumoniae* isolates, penicillin resistance was more common in male patient isolates at 15.4 %, compared with 9.3 % for female patient isolates. The same trend toward increased frequency among male patient isolates was observed with macrolides, cephalosporins, and sulfonamides, with resistance at 35.9 %, 18.5 %, and 53.1 %, respectively, compared with 25.9 %, 10.1 %, and 41.1 % in female patient isolates. Conversely, *Staphylococcus aureus* isolates from female patients were more resistant to penicillins at 43.9 % compared to 29.4 % for male patient isolates. The same pattern was observed for tetracyclines, with 49.8 % of the female patient isolates being resistant versus 32.6 % for male patient isolates. Resistance to sulfonamides was also higher in female patient isolates at 54.6 % than at 35.4 % for male patient isolates.

Among *Enterococcus* isolates, macrolide resistance was significantly higher among female patients at 47.2 % compared to male patient isolates at 22.2 %. Sulfonamide resistance was also seen similarly, and the resistance rates among female and male patient isolates were found to be 72.2 % and 22.2 %, respectively. Resistance in *Streptococcus* species was greater among male patients’ isolates with a rate of 52.9 % and that among female patients’ isolates was 41.2 %. Sulfonamide resistance, however, did not show gender bias and had resistance occurring in 50.0 % of the isolates of male as well as female patients.

### Gram-Negative Bacteria

Gender differences were also noted among Gram-negative bacterial isolates (Figure 6). Resistance against nitrofurans in *Escherichia coli* was greater among female patient isolates, at 57.1 % than among male patient isolates, at 7.1 %. Resistance to cephalosporins was also greater among female patient isolates, at 31.6 % than among male patient isolates, at 15.8 %.

**Figure 6:**
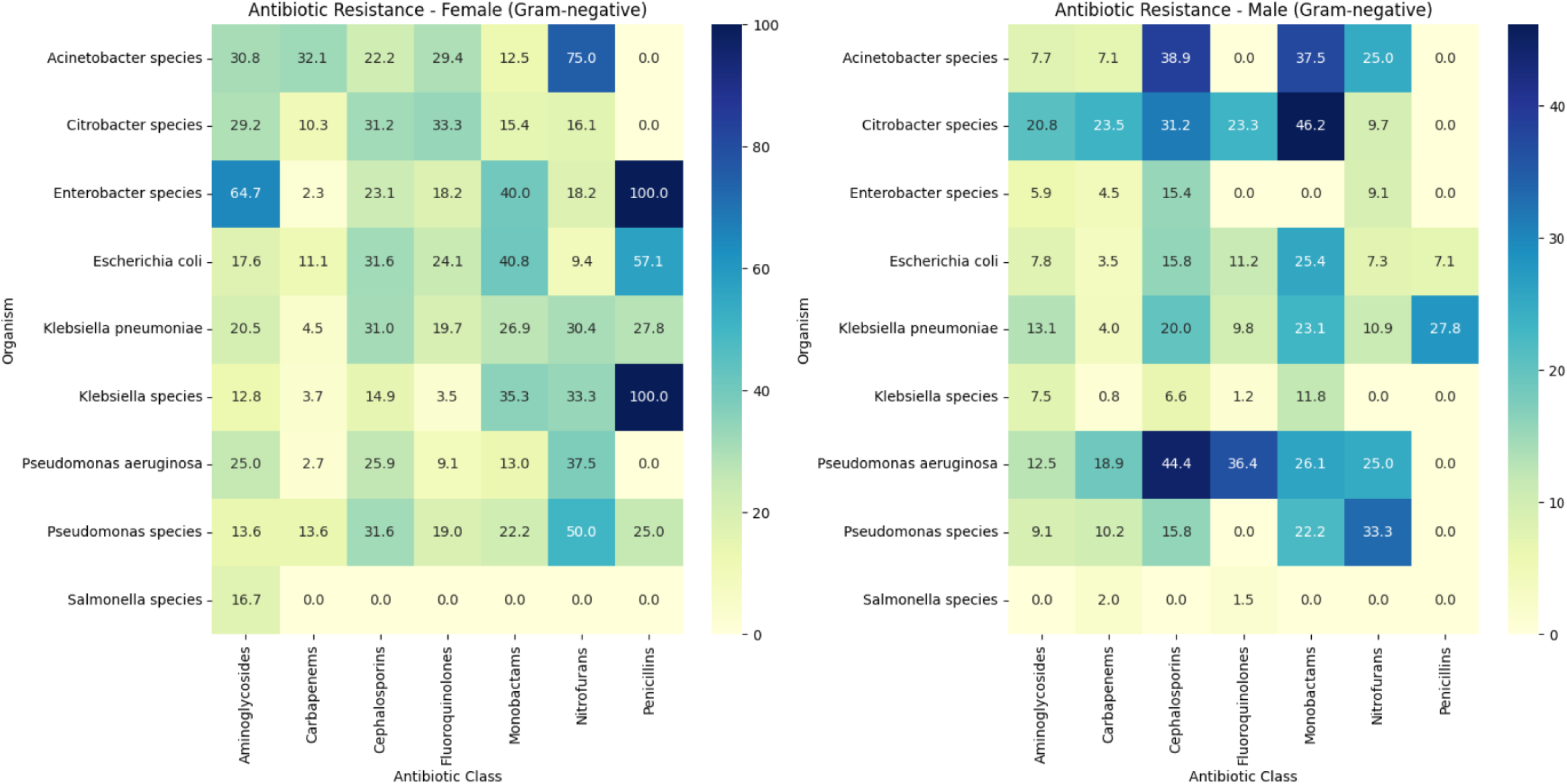
Gender-Specific Resistance in Gram-Negative Organisms

*Klebsiella pneumoniae* also exhibited the same pattern, with cephalosporin resistance at 31.0 % in female patients’ isolates and 20.0 % in male patients’ isolates. Fluoroquinolone resistance was also higher in female-associated isolates, which was at 19.7 %, compared to male-associated isolates at 9.8 %.

*Enterobacter* species isolates from female patients were far more resistant to aminoglycosides, with 64.7 % of the isolates resistant, compared to 5.9 % of male patient isolates. Penicillin resistance was present in all female patient isolates, with no resistance present in male patient isolates. Cephalosporin resistance was more prevalent in female patient isolates, at 23.1 %, compared to 15.4 % for male patient isolates.

Among *Pseudomonas aeruginosa* isolates, fluoroquinolone resistance was more prevalent in male patient isolates, where 44.4 % were resistant, while 37.5 % were resistant among female patient isolates. Cephalosporin resistance was also similar, with resistance standing at 36.4 % among male patient isolates and a lower rate of resistance, 25.9 %, among female patient isolates. In *Acinetobacter spp*, carbapenem resistance was found more often in female patients’ isolates (32.1%) while among male patients’ isolates, resistance was 7.1 %.

The Wilcoxon statistic value of 991.000 and a p-value of 0.00009, indicates a statistically significant difference between the resistance percentages of the two groups. The median percentage of resistance in female patient isolates was 23.08 percent and in male patient isolates was 12.50 percent, with a median difference of 6.67 percent confirming the gender-specific antimicrobial trends statistically.

### Patterns of Multidrug Resistance (MDR) among Bacterial Isolates by Sex

Multidrug-resistant (MDR) isolates were seen more frequently in bacterial isolates from female patients (Figure 7). A total of 2,718 MDR isolates were identified, accounting for 65.18% of all bacterial isolates. The prevalence of MDR isolates differed among bacterial species, and the majority of organisms had a higher rate of MDR isolates among female patients, except *Pseudomonas aeruginosa*. *Escherichia coli* was among the highest in MDR loads in Gram-negative bacteria, with nearly 70 % of the MDR isolates being from female patients. *Klebsiella* species also had more than 65 % of the MDR isolates from female patients. *Enterobacter* species also reported more than 50 % of MDR isolates to be found among female patients, whereas *Acinetobacter* species reported quite an even distribution with about 40 % of MDR isolates being from male patients. For *Pseudomonas aeruginosa*, MDR isolates were found more frequently among male patients, with around 55 % of MDR isolates found among male patients and the remaining 45 % found among female patients.

**Figure 7:**
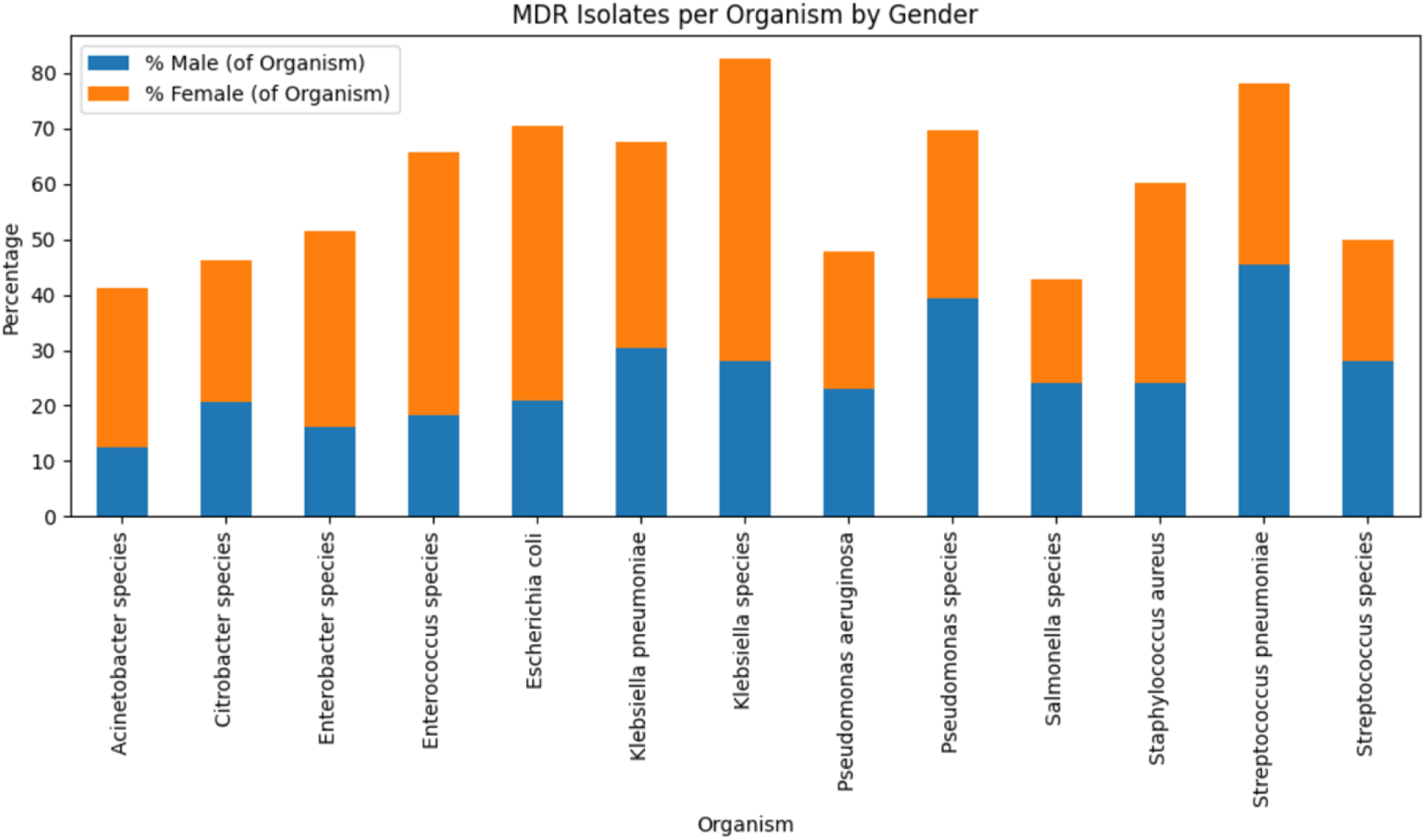
Gender Distribution of Multidrug-Resistant (MDR) Isolates Across Bacterial Species

Among Gram-positive organisms, *Streptococcus pneumoniae* carried over 50 %of its MDR burden in female patient isolates, and *Staphylococcus aureus* carried approximately 60 % of the MDR isolates from female patients. Even *Enterococcus* species contained a majority of MDR isolates from female patients.

The value for the chi-squared statistic of *Escherichia coli* was 6.13 with a p-value of 0.0133, while that of *Staphylococcus aureus* was 9.537 with a p-value of 0.002, showing that MDR isolates for these pathogens were not distributed equally between genders. There was a statistical association between gender and MDR distribution in *Escherichia coli*, odds ratio of 1.66 with a p-value of 0.0118, and in *Staphylococcus aureus*, odds ratio of 0.73 with a p-value of 0.0018. No statistically significant gender differences were noted in the other bacterial species.

## DISCUSSION

The findings of this study highlight the critical role of gender in shaping antimicrobial resistance patterns in Sub-Saharan Africa. With over 92% of bacterial isolates exhibiting resistance to at least one antibiotic and 65% meeting the definition of multidrug resistance (MDR), the data reflect a heavy burden of AMR at Mbarara Regional Referral Hospital. More importantly, the analysis uncovers pronounced and statistically significant gender disparities, with female-derived isolates exhibiting consistently higher resistance rates across key antibiotic classes, particularly beta-lactams, fluoroquinolones, and aminoglycosides.

These disparities align with prior research indicating that women are frequently exposed to broad-spectrum antibiotics, particularly in the management of recurrent urinary tract infections and reproductive health conditions including bacterial vaginosis, pelvic inflammatory disease, and post-surgical prophylaxis (13–17).

UTIs are among the most common bacterial infections globally, and women have a higher predisposition due to anatomical factors such as a shorter urethra, which facilitates bacterial entry into the bladder (18). Additionally, pregnancy and the use of contraceptives have been associated with an increased risk of UTIs in women (19–21). Pregnancy induces physiological changes, including hormonal shifts and urinary stasis due to uterine pressure on the bladder, which predisposes expectant mothers to UTIs (21). Similarly, contraceptives can alter vaginal and urinary tract microbiota, reducing protective normal flora such as lactobacilli and facilitating bacterial colonization, thereby increasing the risk of UTIs (20, 22).

In LMICs like Uganda, where diagnostic capacity is often limited, the frequent recurrence of UTIs among women is also more likely to be prescribed antibiotics empirically amplifying selective pressure and resistance, particularly *Escherichia coli*, the primary UTI pathogen (7, 23). Additionally, *Staphylococcus aureus* is frequently implicated in skin and soft tissue infections, potentially linked to antibiotic use for reproductive and postnatal infections among women (24, 25).

This study found significantly higher MDR rates among *E. coli* and *S. aureus* isolates from female patients, further supporting the hypothesis that antibiotic exposure patterns play a critical role in resistance development thereby echoing global findings on the feminization of AMR burden in community-acquired infections (5, 26)

Conversely, higher MDR prevalence in male-derived *Pseudomonas aeruginosa* isolates likely reflects gendered exposure to nosocomial pathogens, as men are often more represented in ICU admissions and invasive procedures associated with healthcare-acquired infections (27, 28). Additionally, men are more likely to only present themselves in hospital with chronic conditions such as diabetes, chronic obstructive pulmonary disease (COPD), and other immunosuppressive diseases (29), which usually require prolonged hospitalization and long antibiotic regimens, further increasing the risk of MDR development. Men also exhibit higher rates of smoking and alcohol consumption, which have been associated with impaired immune function and an increased risk of hospital-acquired infections (29). These lifestyle factors, combined with higher rates of occupational exposure to environmental pathogens in industries such as construction, mining, and agriculture, may contribute to increased AMR burden in male patients (29, 30). Furthermore, adherence to treatment regimens tends to be lower among men, potentially leading to the selection of resistant bacterial populations (31, 32).

These differences emphasize that AMR is not only a microbiological issue but also a sociostructural one, shaped by gendered access to care, prescribing patterns, and infection risks.

The World Health Organization’s Global Antimicrobial Resistance Surveillance System (GLASS) encourages the collection of sex-disaggregated data (33), yet implementation remains uneven in many LMICs. Additionally, public health messaging and stewardship campaigns should consider gender in their design and delivery. For example, embedding AMR education in reproductive health services and antenatal clinics could help reduce inappropriate antibiotic use among women (34). At the same time, improving hospital infection prevention for male patients—particularly in high-risk units—may reduce exposure to resistant nosocomial organisms (35).

As a retrospective study, it relies on existing hospital records, which may be subject to missing data. Additionally, the lack of molecular characterization of resistance mechanisms limits deeper insights into genetic determinants of AMR disparities. Future research should explore the molecular mechanisms underlying observed gender differences, the role of self-medication, and intersectional factors such as age, socioeconomic status, and comorbidities contributing to gender-based AMR trends. Understanding these dynamics can inform targeted interventions that improve both clinical outcomes and health system equity.

## Conclusion

This decade-long retrospective study provides robust microbiological evidence of gender disparities in AMR and MDR in a Ugandan tertiary care setting. These patterns reflect the complex interplay between biological vulnerability, antibiotic exposure, and healthcare access.

This study provides significant microbiological evidence of gender differences in AMR and MDR in Uganda, emphasizing the importance of routine gender-based metrics in AMR surveillance. The findings argue for including gender-sensitive measures in Uganda’s national AMR action plan and global AMR response strategy. Addressing gender inequities through targeted antimicrobial stewardship policies and increased diagnostic capacity will be essential to achieving fair and effective AMR mitigation policies in LMICs

## Data Availability

The datasets used and analyzed during the current study are available from the corresponding author upon reasonable request.

## Ethical Clearance

The study is under a big project "AMR Surveillance, Modelling Antimicrobial Pharmaceutical Needs and PCR/Herbal/Biotechnological Products Development" in Uganda. Approval was obtained from Busitema University Faculty of Health Sciences REC with REC number BUFHS-2022-15. This study did not interface with patients as the clinical isolates were obtained from patients under routine hospital procedures.

### Author contribution

All authors contributed equally to the conception, design, methodology, and writing of this manuscript.

